# Deep Learning–Based Early Detection of Major Adverse Cerebral Injuries in Cardiothoracic and Vascular Surgery

**DOI:** 10.1101/2025.01.10.25320368

**Authors:** Dongjoon Yoo, Yunwon Tae, Kyungjae Cho, Hyunggon Je, Dohyung Kim, Bongsoo Son, Minho Ju, Cheehoon Lee, Sangsu Lee, Taehwa Kim, Woo Hyun Cho, Young A. Kim, Narae Lee, Sung-Ho Ahn

## Abstract

**Background:** Despite advances in central nervous system (CNS)-protective anesthetic and surgical strategies, perioperative stroke remains a significant concern in high-risk cardiothoracic and vascular surgery (CTVS). Early detection, facilitating timely and prompt intervention, is often hindered by sedation and mechanical ventilation (MV) in the immediate postoperative period. This study aimed to develop and validate a deep learning (DL)-based artificial intelligence (AI) program for early detection of severe, surgery-related major adverse cerebral injury (sMACI), encompassing fatal CNS and systemic insults in high-risk CTVS patients.

**Methods:** We retrospectively analyzed data from 4,455 patients who underwent seven types of CTVS (2010–2021), requiring postoperative ICU admission and ongoing MV. Continuous vital signs (heart rate, blood pressures, respiratory rate, pulse oximetry saturation, temperature) were extracted from the operating room (OR) and intensive care unit (ICU), along with demographic and laboratory data. sMACI was defined as significant postoperative CNS injury (modified Rankin Scale ≥3 at 1 month) or 1-month mortality. Two-tier DL models were constructed: Model 1 using ICU data alone, and Model 2 integrating pre-ICU and ICU data. Performance in detecting sMACI within 24 hours of ICU admission was assessed using the area under the receiver operating characteristic curve (AUROC) and precision-recall curve (AUPRC).

**Results:** Among 4,455 patients, 5% experienced sMACI. Model 1 achieved an AUROC of 0.809 (95% CI: 0.759–0.858) and an AUPRC of 0.275 (0.195–0.375). Model 2 showed improved detection (AUROC 0.826 [0.781–0.871]; AUPRC 0.322 [0.233–0.423]). Both models outperformed conventional early warning scores and other machine learning algorithms, demonstrating robust performance as early as 4 hours after ICU admission. Key contributors included systolic blood pressure, heart rate, diastolic blood pressure, mean arterial pressure, and pulse oximetry saturation.

**Conclusions:** A DL-based AI program leveraging continuous vital signs enables effective early detection of severe surgery-related CNS and systemic injury in high-risk CTVS patients, outperforming established scoring systems and other machine learning approaches.

## Introduction

Perioperative stroke (PS), defined as a cerebrovascular event occurring during or shortly after surgery, significantly affects patient outcomes, leading to prolonged hospital stays, long-term disabilities, and mortality, as well as increased socio-economic burdens.^1, 2^ Despite advancements in anesthetic and surgical techniques that have reduced perioperative mortality and myocardial infarctions, the incidence of PS remains substantial at 1-5% in coronary artery bypass grafting and valve surgery,^3, 4^ 2-10% in aortic surgeries, and 2-5% in major vascular and thoracic procedures.^5, 6^

Guidelines for cardiac and non-cardiac surgeries emphasize strategies to reduce perioperative central nervous system (CNS) injury risk,^7^ such as identifying high-risk patients through comprehensive preoperative assessments, optimizing medical risk factor management, and maintaining stable hemodynamics throughout the perioperative phases, to effectively implement CNS-protective measures.^8–12^ Additionally, early recognition and prompt diagnosis of CNS damage are crucial, as timely interventions, such as imaging-based intra-arterial thrombectomy (recently permitted within 24 hours or even in unclear onset cases) for emergent large-vessel occlusion strokes,^13, 14^ and urgent management of brain edema and intracranial hypertension, particularly in cases of severe CNS insults,^15, 16^ can significantly mitigate its severity.

However, early detection of surgery-related CNS injury remains challenging, particularly during the immediate post-operative intensive care unit (ICU) stay.^17, 18^ Neurological assessments are often confounded by anesthetics, sedatives, muscle relaxants, analgesics, and ongoing mechanical ventilation (MV), frequently delaying diagnosis until MV is discontinued after the weaning process.^19^

In this study, we hypothesized that early surgery-related CNS injury could be detected through machine learning (ML) analysis of real-time biosignals, such as continuous vital signs routinely monitored in the ICU and operating room (OR).^20, 21^ These signals may capture the complex brain-heart interaction, ^22^ whereby CNS damage alters systemic physiology via neurogenic autonomic dysregulation.^23, 24^ We aimed to develop a deep learning (DL)-based artificial intelligence (AI) program capable for identifying subtle changes in vital signs, derived from neurogenic stress, that indicate impending CNS complications in high-risk cardiothoracic and vascular surgery (CTVS) patients during the immediate post-operative ICU stay.

## Methods

### Study design

This single-center retrospective study developed a DL-based AI program to detect early surgery-related CNS damage. Although DL can achieve high accuracy and automatically learn complex features from large datasets, it presents challenges, including limited interpretability and potential overfitting.^25^ To address these, we implemented a systematic model development strategy focused on interpretability and clinical applicability.

Initially, we introduced the concept of surgery-related major adverse cerebral injuries (sMACI) and a fixed reference time point (24 hours after ICU admission) to evaluate our model. We hypothesized that vital sign instability during the first 24 hours in the ICU may reflect concurrent CNS and systemic injury severity. Using sMACI as the primary outcome, we aimed to determine whether vital sign instability correlates with early detection capability (Figure 1A).

**Figure 1.**
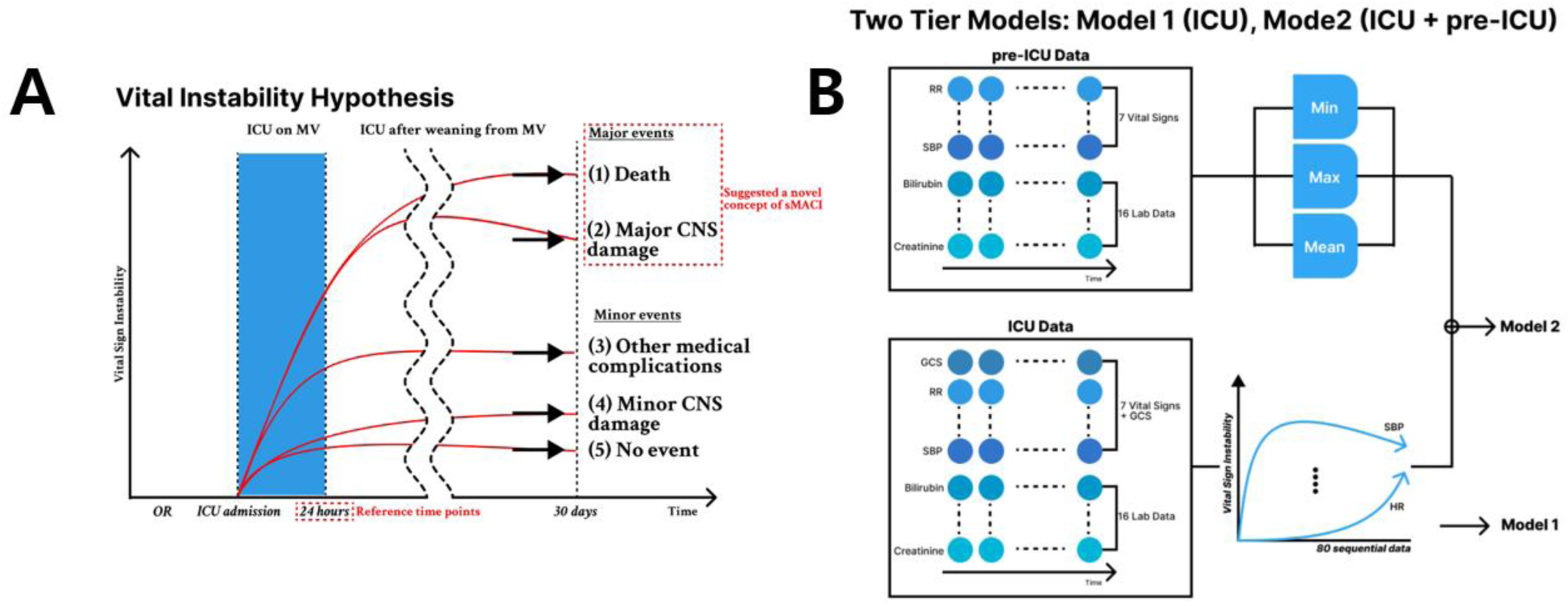
Study hypothesis and model design. CNS, central nervous system; ICU, intensive care unit; MV, mechanical ventilator; OR, operative room; sMACI, surgery-related major adverse cerebral injuries

To ensure versatility, we included seven representative CTVS procedures and developed a standardized dataset encompassing both ICU and pre-ICU (intraoperative and preoperative) data. We devised a two-tier model structure: Model 1 used ICU data only, while Model 2 incorporated both pre-ICU and ICU data. This design allowed us to examine whether early vital sign data (pre-ICU) improve detection accuracy and generalizability. We also assessed the feasibility of detection earlier than the standard 24-hour reference point, such as within the initial hours of MV therapy (Figure 1B).

### Study population

We included consecutive CTVS-ICU patients admitted postoperatively at Pusan National University Yangsan Hospital (PNUYH) from March 2010 to March 2021. Inclusion criteria were patients requiring intensive postoperative monitoring without recovery-room extubation. The Institutional Review Board of PNUYH approved this study (IRB No. 04-2022-001) and waived the requirement for individual consent. The study followed TRIPOD guidelines. Of 6,162 cases, 1,707 were excluded due to repeated surgeries during the same admission, preexisting CNS damage, organ transplantation, or missing data. The final cohort comprised 4,455 patient records from seven different CTVS procedures (Supplemental Figure 1).

### Data collection and preprocessing

We extracted electronic health record (EHR) data, including vital signs, demographic information, surgical details, and laboratory results, up to 24 hours post-ICU admission. Key vital signs included heart rate (HR), systolic/diastolic/mean arterial blood pressures (SBP/DBP/MAP), respiratory rate (RR), pulse oximetry saturation (SpO2), and body temperature (BT). Glasgow Coma Scale (GCS) scores evaluated consciousness.^26^ We included demographic data, surgical variables, and 16 laboratory tests.

### Outcome assessments

The primary outcome, sMACI, was defined as significant early-phase postoperative CNS injury or fatal systemic complications resulting in death within one month. Major CNS damage was identified by severe ischemic or hypoxic brain injury during or at the end of MV weaning within 7 days of surgery, accompanied by substantial neurological deficits (modified Rankin scale ≥3 at 1 month).^27^ One-month mortality indicated fatal systemic damage. An independent neurologist reviewed EHR and brain imaging. The control group included minor CNS injuries (e.g., minor stroke, isolated seizures) or other nonurgent medical complications, as well as patients without events.

### Model development

We used a four-step model development approach (Figure 2):

1. Data preparation: We selected 4,455 of 6,162 cases for analysis (2010–2017 as development set; 2018–2021 as validation set). Based on established operative risk systems, we included 4 demographic/surgical variables, 7 vital signs, GCS scores, and 16 laboratory results. Pre-ICU features excluded GCS scores. Invasive measurements were prioritized.
2. Data pre-processing and missing value imputation: Values outside valid ranges (Supplemental Table 1) were treated as missing. Missing values arose from EHR asynchrony, mitigated by hourly or daily aggregation. Non-numeric entries were treated as missing. A heatmap showing the missing data rate is in the Supplement Figure 2. Missing values were forward filled. Variables were zero-padded for equal sequence length. Vital signs and laboratory values were z-score normalized. Pre-ICU data were summarized by minimum, maximum, and mean values.
3. Model development and validation: The model comprised (1) ICU feature-embedding module: one fully connected network (FCN) and normalization layer processing 80 sequential data points; (2) Sequential characteristics module: three bidirectional long short-term memory networks (LSTMs)^28^ to aggregate 80 sequential data points; (3) Demographic- embedding modules: static demographic features processed by an FCN; (4) Pre-ICU feature-embedding module: summarized intraoperative and preoperative vital signs processed by an FCN. Dropout regularization was applied.^29^ This technique enhanced the model’s robustness and generalization capabilities, aligning with methodologies established in previous studies.^30^ Model 2 used ICU and pre-ICU features; Model 1 used only ICU data (demographic included in all).
4. Evaluation: We computed the area under the receiver operating characteristic curve (AUROC), area under the precision-recall curve (AUPRC), sensitivity, specificity, positive predictive value (PPV), negative predictive value (NPV), and F-measure at 24 hours post-ICU admission. For patients staying <24 hours, evaluations were postponed until 24 hours to ensure uniformity.

**Figure 2.**
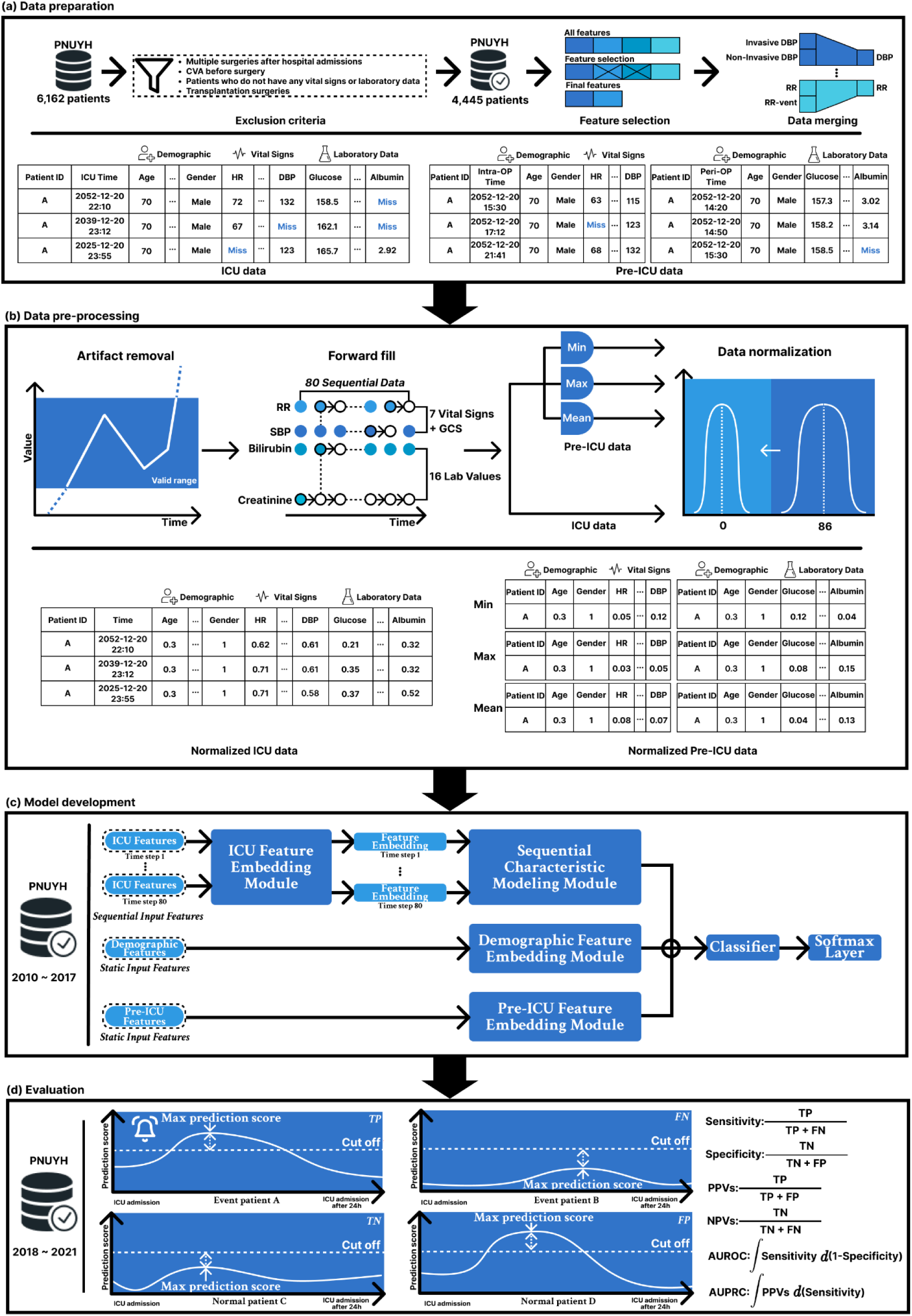
Development process and validation overview. AUPRC, area under the precision-recall curve; Miss, missing values; AUROC, area under the receiver operating characteristic curve; CVA, cerebrovascular accident; DBP, diastolic blood pressure; FN, false negative; FP, false positive; GCS, Glasgow Coma Scale; HR, heart rate; ICU, intensive care unit; ID, identity document; Max, maximum; Min, minimum; NPVs, negative predictive values; OR, operative room; PNUYH, Pusan National University Yangsan Hospital; PPVs, positive predictive values; RR, respiratory rate; SBP, systolic blood pressure; TN, true negative; TP, true positive;

### Model performance and evaluation

We compared Models 1 and 2 against established scoring systems such as the National Early Warning Score (NEWS) and the Modified Early Warning Score (MEWS), which were designed to assess illness severity,^31^ and other ML algorithms, such as random forest (RF), Light Gradient Boosting Machine (LightGBM), and logistic regression (LR), commonly used in predicting critical events following surgical or medical procedures.^32–35^ Statistical significance was assessed using DeLong’s test. We evaluated performance at various time intervals relative to the 24-hour mark.

### Systemic verification

We verified sMACI validity as the primary outcome and tested the hypothesis that vital sign instability drives model performance. We examined distributions of key vital signs in ICU and pre-ICU settings, refined our hypothesis, and progressively incorporated pre-ICU data.

Shapley Additive Explanations (SHAP) were used to clarify input variable contributions to the model’s detection power.^36^

### Statistical analysis

Statistical differences between development and validation were evaluated using Welch’s t-test and the chi-square test for continuous and categorical variables, respectively. Statistical tests were computed using Python (v3.8.5) using sklearn (v0.24). Statistical significance was tested using DeLong’s test (p<0.001) in R (v3.6.3) (http://www.r-project.org). Confidence intervals (CI) were computed using 1,000 bootstraps.

## Results

### Cohort characteristics

From 4,455 eligible cases over 11 years, we identified 2,773,281 ICU data points (mostly hourly recorded vital signs) and 1,130,756 pre-ICU data points (mostly every 10 minutes recorded vital signs in OR) (Supplemental Table 1).

The mean ICU stay was 67.97±193.44 hours (median: 24.32 [21, 43.1] hours), and mean OR stay was 3.99±2.26 hours (median: 2.5 [3.5, 4.95] hours). About 5% of patients developed sMACI (Supplemental Figure 3). Demographics and clinical features differed between development and validation sets in surgery types and some labs (Supplemental Table 2).

### Detection performance

Model 1 (ICU data only) achieved an AUROC of 0.809 (95% CI: 0.759–0.858) and AUPRC of 0.275 (0.195–0.375) in Table 1, outperforming NEWS, MEWS, and other ML algorithms (Figure 3A, Supplemental Table 3).

**Figure 3.**
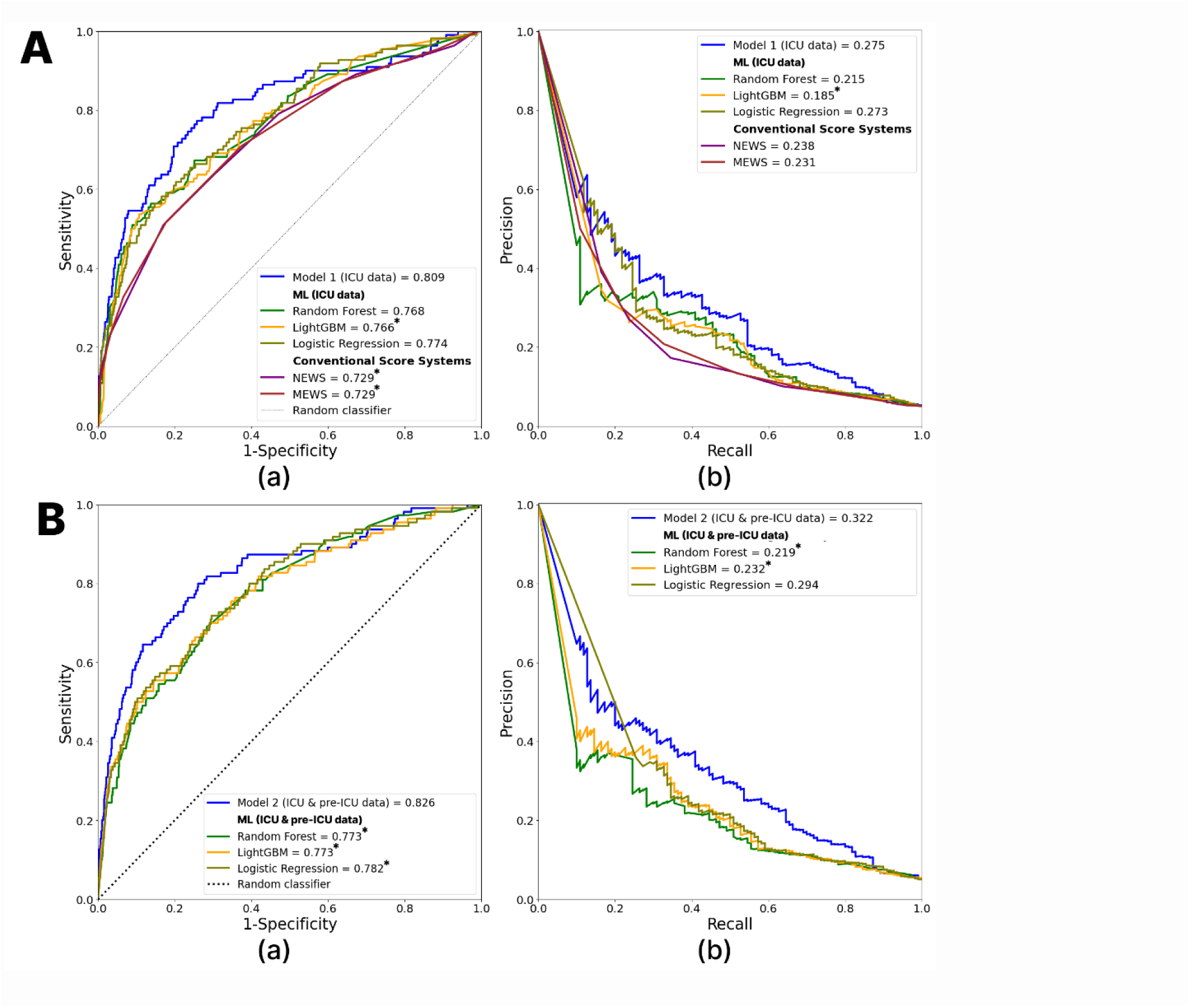
AUROCs and AUPRCs for early detection of sMACI in Model 1 (A) and Model 2 (B). *p<0.05, DeLong’s test for AUROC or paired t-test for AUPRC were used to determine the significance of the observed differences between Models 1 and 2. AUPRC, area under the precision-recall curve; AUROC, area under the receiver operating characteristic curves; GBM, Gradient Boosting Machine; ICU, intensive care unit; LR, logistic regression; MEWS, Modified Early Warning Score; ML, machine learning; NEWS, National Early Warning Score; RF, random forest; sMACI, surgery-related major adverse cerebral injuries.

**Table 1.**
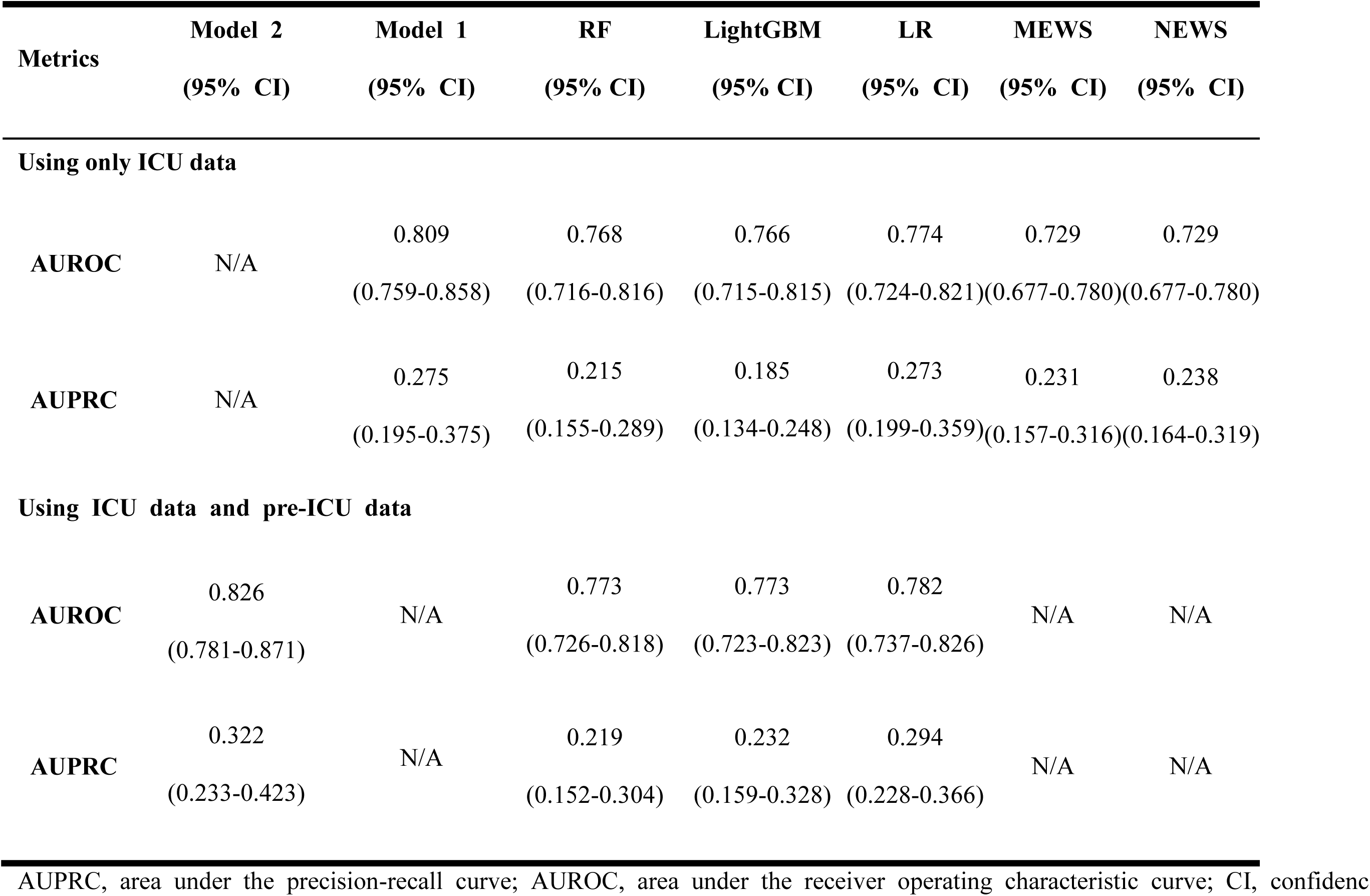
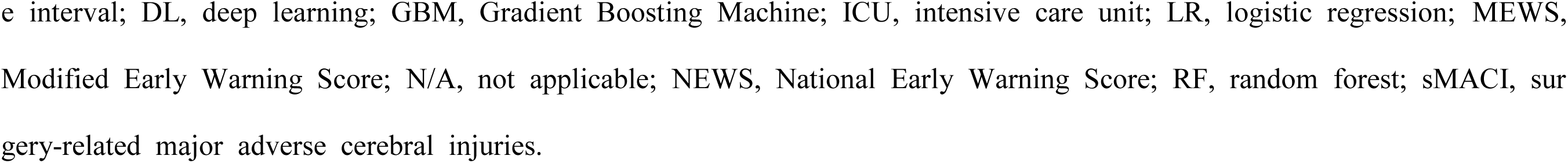
Comparison of DL-based and conventional scoring systems for detecting sMACI within 24 h of ICU admission.

| **Metrics** | **Model 2**<br>**(95% CI)** | **Model 1**<br>**(95% CI)** | **RF**<br>**(95% CI)** | **LightGBM**<br>**(95% CI)** | **LR**<br>**(95% CI)** | **MEWS**<br>**(95% CI)** | **NEWS**<br>**(95% CI)** |
| --- | --- | --- | --- | --- | --- | --- | --- |
| **Using only ICU data** |  |  |  |  |  |  |  |
| **AUROC** | N/A | 0.809<br>(0.759-0.858) | 0.768<br>(0.716-0.816) | 0.766<br>(0.715-0.815) | 0.774<br>(0.724-0.821) | 0.729<br>(0.677-0.780) | 0.729<br>(0.677-0.780) |
| **AUPRC** | N/A | 0.275<br>(0.195-0.375) | 0.215<br>(0.155-0.289) | 0.185<br>(0.134-0.248) | 0.273<br>(0.199-0.359) | 0.231<br>(0.157-0.316) | 0.238<br>(0.164-0.319) |
| **Using ICU data and pre-ICU data** |  |  |  |  |  |  |  |
| **AUROC** | 0.826<br>(0.781-0.871) | N/A | 0.773<br>(0.726-0.818) | 0.773<br>(0.723-0.823) | 0.782<br>(0.737-0.826) | N/A | N/A |
| **AUPRC** | 0.322<br>(0.233-0.423) | N/A | 0.219<br>(0.152-0.304) | 0.232<br>(0.159-0.328) | 0.294<br>(0.228-0.366) | N/A | N/A |

AUPRC, area under the precision-recall curve; AUROC, area under the receiver operating characteristic curve; CI, confidence
e interval; DL, deep learning; GBM, Gradient Boosting Machine; ICU, intensive care unit; LR, logistic regression; MEWS, Modified Early Warning Score; N/A, not applicable; NEWS, National Early Warning Score; RF, random forest; sMACI, surgery-related major adverse cerebral injuries.

Model 2 (ICU and pre-ICU data) improved to an AUROC of 0.826 (0.781–0.871) and AUPRC of 0.322 (0.233–0.423) in Table 1, surpassing Model 1 and other benchmark methods (Figure 3B, Supplemental Table 3).

### Threshold for earlies detection time

Evaluations of performance of Model 1 and 2 at various intervals showed robust performance relative to the reference time of 24 hours of ICU stay (Supplemental Table 4).

Model 1 demonstrated robust performance, with an AUROC ranging from 0.809 (95% CI: 0.759–0.858) at 24 hours to 0.797 (95% CI: 0.744–0.849) at 4 hours after ICU admission, consistently outperforming NEWS, MEWS, and other ML algorithms (Figure 4A, Supplemental Table 5).

**Figure 4.**
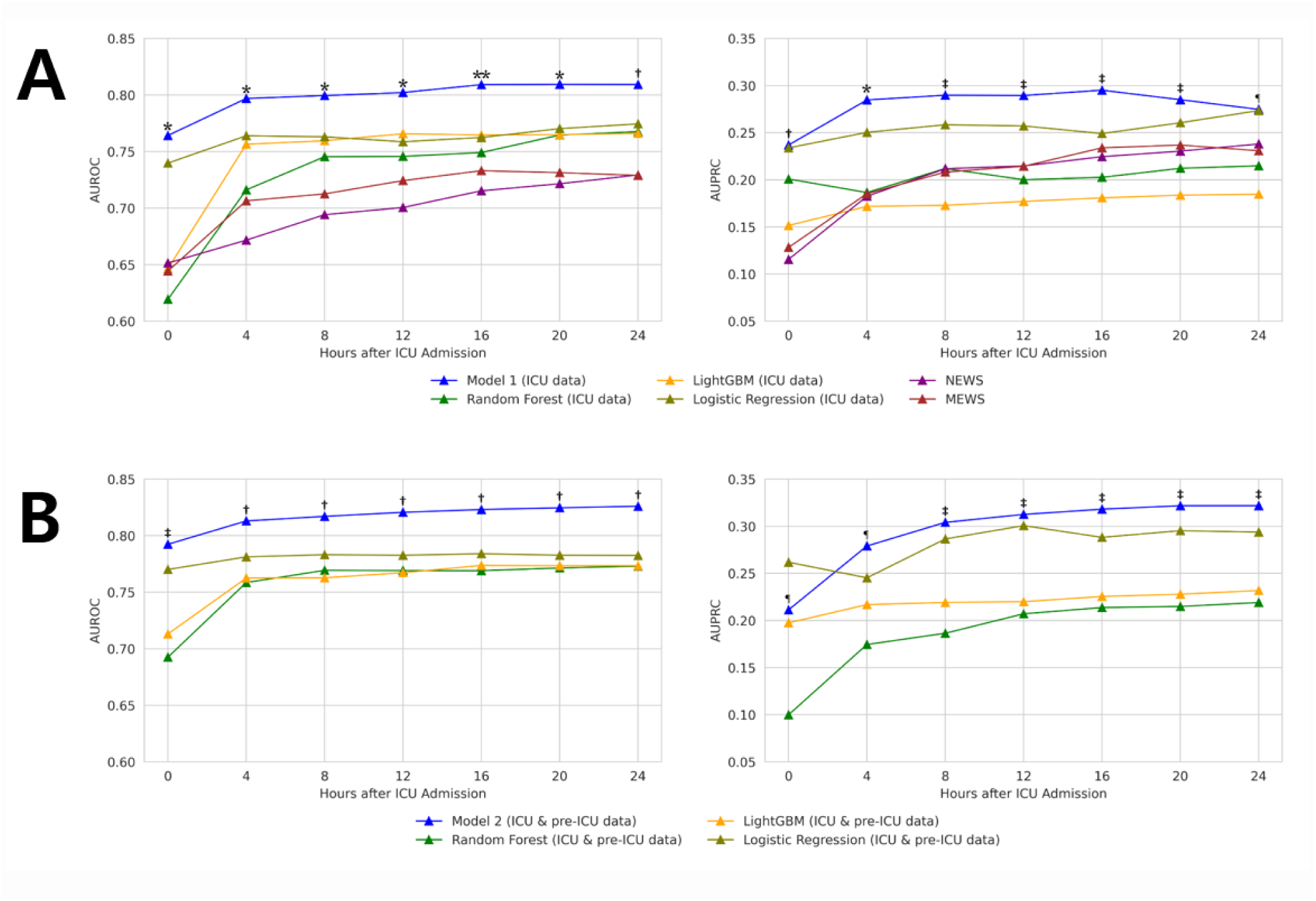
Early detection of sMACI at different time windows in Model 1 (A) and Model 2 (B). **p<0.05 across 5 models, *p<0.05 across 4 models, †p<0.05 across 3 models, ‡p<0.05 across 2 models, ¶p<0.05 for 1 model, as assessed by DeLong’s test for AUROC or paired t-test for AUPRC to determined he significance of the observed differences between Models 1 or 2. AUPRC, area under the precision-recall curve; AUROC, area under the receiver operating characteristic curves; GBM, Gradient Boosting Machine; ICU, intensive care unit; LR, logistic regression; MEWS, Modified Early Warning Score; NEWS, National Early Warning Score; RF, random forest; sMACI, surgery-related major adverse cerebral injuries.

Model 2 showed similar strength, with an AUROC ranging from 0.826 (95% CI: 0.781–0.871) at 24 hours to 0.813 (95% CI: 0.768–0.859) at 4 hours after ICU admission, consistently superiority to other benchmark methods (Figure 4B, Supplemental Table 5).

### Systemic verification

Distribution of key vital signs differed by presence of sMACI (Supplemental Figure 4A) and varied with more specific events (Supplemental Figure 4B) during the ICU stay.

Additionally, vital sign distributions remained stable across CTVS procedures in the ICU setting (Supplemental Figure 5A), while pre-ICU distributions varied by CTVS procedure (Supplemental Figure 5B).

SHAP analysis identified SBP, HR, DBP, MBP, SpO2, GCS components, and summarized pre-ICU vital signs and surgical parameters as key contributors (Figure 5).

## Discussion

### 1) Main summary

In this large-scale study of individual-level data, which included over 2.7 million ICU data points and 1.1 million pre-ICU data points, we developed and validated an AI-assisted early detection program using DL algorithms to detect sMACI in patients undergoing high-risk CTVS within the first 24 hours of their postoperative ICU stay.

**Figure 5.**
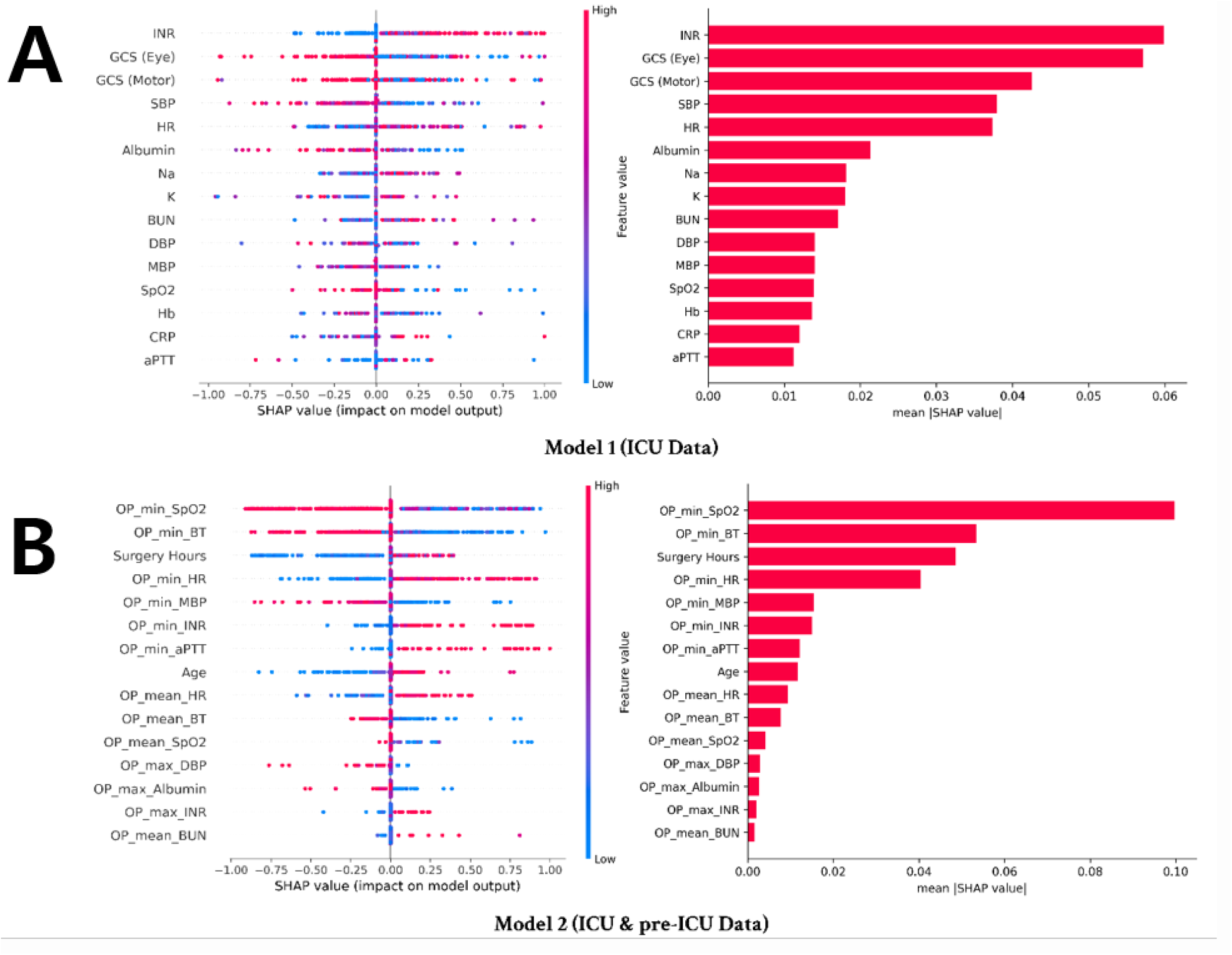
Feature importance of 950 random samples of vital signs, laboratory data, and demographic features in Model 1 (A) and Model 2 (B). Each dot on the graph represents a randomly selected patient at a specific time during their ICU stay, with red denoting a high feature value and blue a low feature value. The left side of the plot represents the top contributing features, arranged from top (highest) to bottom (lowest). The color of each dot represents the feature values, and the x-axis represents the Shapley values. Negative and positive Shapley values indicate whether a feature contributes to producing low or high-risk scores, respectively. The right side of the plot shows the absolute mean Shapley values, summarizing the importance of the overall feature. aPTT, activated partial thromboplastin time; BT, body temperature; BUN, blood urea nitrogen; CRP, C-reactive protein; DBP, diastolic blood pressure; GCS, Glasgow Coma Scale; HR, heart rate; Hb, hemoglobin; ICU, intensive care unit; INR, international normalized ratio; K, potassium; MBP, mean blood pressure; max, maximum; min, minimum; Na, sodium; OP, operation; SBP, systolic blood pressure; SHAP, Shapley Additive Explanations; SpO_2_, oxygen saturation.

Our structured approach aimed to enhance both interpretability and versatility of DL models, addressing their inherent limitations to increase clinical utility.^37^ We introduced the novel concept of sMACI as a primary outcome measure and defined reference time points to capture dynamic interactions between degree of vital sign instability and stage of MV therapy. We also implemented a two-tier model: Model 1 utilizing ICU data alone and Model 2 incorporating both pre-ICU and ICU data. This design allowed us to investigate whether the inclusion of pre-ICU vital sign data improves detection accuracy and highlights the critical role of early physiological signals, confirming our hypothesis. By incorporating comprehensive ICU and pre-ICU data from diverse CTVS procedures, this approach ensures adaptability across different clinical settings and data extraction methods. Additionally, we conducted systematic verification to confirm our study hypothesis and model design.

### 2) Clinical relevance

Our DL-based approach, leveraging continuous biosignals, outperformed traditional scoring systems (e.g., NEWS and MEWS) and other ML algorithms, demonstrating reliable detection as early as 4 hours after ICU admission.

Early identification of sMACI is crucial, given the high risk of severe CNS and systemic damage following CTVS, despite advancements in perioperative care. Current guidelines and CNS treatment protocols emphasize the importance of early detection to enable timely interventions. For example, imaging-guided endovascular therapy (including intra-arterial thrombectomy) now extends the therapeutic window to 24 hours post-onset, or even longer if the onset time is unclear.^38, 39^ Such strategies are particularly relevant during the perioperative period of CTVS, where prompt intervention can significantly influence outcomes.

Furthermore, proactive management of brain edema and intracranial pressure is essential to mitigate severe PS or hypoxic-ischemic brain damage,^16^ as well as complications like post-stroke or post-hypoxic intractable seizures.^40^

Our findings underscore the potential of AI to improve patient outcomes through real-time, continuous assessments integrated into routine care. While direct comparisons of sMACI incidence to previous studies are limited by different time point definitions, our observed rate of approximately 5% aligns with known complication rates.^3–6^

### 3) Development process of two-tier models

We systematically developed the model to address interpretability challenges inherent to DL algorithms.^41^ Initially, we formulated a study hypothesis that vital sign instability would reflect both the severity of the damage and its relationship to the timing of MV therapy within the first 24 hours of ICU admission. Building on this concept, we introduced sMACI as the primary outcome measure, emphasizing its prognostic importance and the need for urgent intervention, as well as its presumed association with dynamic patterns of vital sign instability.

Furthermore, to enhance generalizability, we targeted a comprehensive CTVS-ICU cohort encompassing various CTVS procedures and utilized a standardized dataset that included both ICU and pre-ICU data. This led to the development of a tailored two-tier AI program incorporating variables from the pre-ICU and ICU periods up to the first postoperative day.

This approach offers two key advantages: (1) enabling early detection for timely interventions before MV weaning, and (2) reducing confounding factors from delayed-onset complications, such as infections, electrolyte imbalances, nutritional issues, or drug interactions during extended ICU stays.^42^ Ultimately, our methods support the scalability and applicability of AI-driven approaches across diverse patient populations and clinical environments.

### 4) Performance comparison and systemic verification

By evaluating DL models using AUROC and AUPRC, we demonstrated their superiority over static scoring systems (NEWS, MEWS) and other ML algorithms for early sMACI detection. Continuous biosignal monitoring was crucial for identifying subtle changes in critically ill patients and enabling timely interventions. DL-based models, incorporating deep neural networks (DNNs) and bidirectional LSTM networks, improved detection accuracy by handling complex sequential data and temporal dependencies.^43^

Model 2, which integrated data from the OR to ICU admission, further improved early detection capabilities and underscored the value of incorporating pre-ICU patient data. While ICU data appeared consistent, likely due to standardized ICU practices, pre-ICU data varied by CTVS subtype, highlighting the need for procedure-specific adjustments.

Additionally, systematic verification further supported our model’s interpretability and validated the study hypothesis by examining vital sign distributions and employing SHAP analysis to elucidate the influence of specific input variables on model predictions. These findings confirm the central role of continuous monitoring and early intervention based on subtle vital sign changes and enhance clinicians’ confidence in AI-assisted decision-making.

Through these development processes, our two-tier tailored DL architecture is among the first designed to facilitate early detection of surgery-related fatal CNS and systemic damage in high-risk CTVS patients, prioritizing interpretability and versatility. This approach holds potential applicability across a broad range of patient populations and clinical settings, advancing perioperative care and improving patient outcomes.

## Study limitations

This study has several limitations. First, its single-center, retrospective design may introduce selection bias and limit generalizability. Nevertheless, it provides a critical foundation for validating the sMACI concept and guiding future large-scale, prospective, multicenter investigations. We also aimed to develop a model encompassing a broad range of CTVS-ICU procedures, enhancing versatility. Our approach demonstrated balanced or superior AUROC and AUPRC performance across different CTVS procedures (Supplemental Table 6), suggesting that it may be broadly applicable in future multicenter CTVS-ICU settings.

Second, relying on EHR data introduces variability in data quality. Although we employed imputation techniques and expert reviews by stroke specialists and intensivists, inaccuracies may persist. Third, differences in clinical practice, perioperative care, anesthesia protocols, and postoperative management could influence model performance. To mitigate this, we incorporated all relevant variables from the pre-ICU period through the first postoperative ICU day, reducing confounding factors and facilitating early detection. Future multicenter studies with standardized variables may further improve robustness and generalizability.

Fourth, the study focused on early detection of surgery-related CNS damage around the time of MV termination rather than broader predictive capabilities. This narrower focus excluded certain patient groups, such as those with preexisting CNS damage or delayed PS, potentially limiting a more comprehensive assessment of the model’s utility. However, this targeted approach mirrors practical clinical needs, as delayed-onset complications are generally more detectible during subsequent ICU monitoring than during MV or weaning.

## Conclusions

We developed a DL-based, AI-assisted early detection program for identifying severe, surgery-related CNS and systemic damage (sMACI) in high-risk CTVS patients within the first 24 hours of their postoperative ICU stay. By leveraging real-time biosignal analysis, our two-tier DL models, designed to improve both interpretability and versatility while addressing the inherent limitations of DL, demonstrated enhanced detection accuracy compared to conventional scoring systems and other ML algorithms. Crucially, our approach holds the potential for identifying critical damage as early as four hours after ICU admission, enabling timely interventions and ultimately improving patient outcomes.

## Data Availability

N/A

## Acknowledgements

This research was supported by a grant from the Korea Health Technology R&D Project through the Korea Health Industry Development Institute (KHIDI), funded by the Ministry of Health & Welfare (RS-2021-KH114109) and by a National Research Foundation of Korea grant (RS-2023-00213690 to S.H.A.), funded by the Korean government (MSIT).

## DISCLOSURES

We declare no competing interests.

## Notes

### Competing Interest Statement

The authors have declared no competing interest.

### Clinical Trial

N/A

### Author Declarations

The Institutional Review Board of PNUYH approved this study (IRB No. 04-2022-001) and waived the requirement for individual consent.

## REFERENCES

1. Selim M. Perioperative stroke. The New England journal of medicine. 2007;356:706–713

2. Gaudino M, Benesch C, Bakaeen F, DeAnda A, Fremes SE, Glance L, et al. Considerations for reduction of risk of perioperative stroke in adult patients undergoing cardiac and thoracic aortic operations: A scientific statement from the american heart association. Circulation. 2020;142:e193–e209

3. Newman MF, Mathew JP, Grocott HP, Mackensen GB, Monk T, Welsh-Bohmer KA, et al. Central nervous system injury associated with cardiac surgery. *Lancet (London*, England). 2006;368:694–703

4. Roach GW, Kanchuger M, Mangano CM, Newman M, Nussmeier N, Wolman R, et al. Adverse cerebral outcomes after coronary bypass surgery. Multicenter study of perioperative ischemia research group and the ischemia research and education foundation investigators. The New England journal of medicine. 1996;335:1857–1863

5. Jorgensen ME, Torp-Pedersen C, Gislason GH, Jensen PF, Berger SM, Christiansen CB, et al. Time elapsed after ischemic stroke and risk of adverse cardiovascular events and mortality following elective noncardiac surgery. Jama. 2014;312:269–277

6. Ng JL, Chan MT, Gelb AW. Perioperative stroke in noncardiac, nonneurosurgical surgery. Anesthesiology. 2011;115:879–890

7. Benesch C, Glance LG, Derdeyn CP, Fleisher LA, Holloway RG, Messé SR, et al. Perioperative neurological evaluation and management to lower the risk of acute stroke in patients undergoing noncardiac, nonneurological surgery: A scientific statement from the american heart association/american stroke association. Circulation. 2021;143:e923–e946

8. Otto CM, Nishimura RA, Bonow RO, Carabello BA, Erwin JP, 3rd, Gentile F, et al. 2020 acc/aha guideline for the management of patients with valvular heart disease: Executive summary: A report of the american college of cardiology/american heart association joint committee on clinical practice guidelines. Circulation. 2021;143:e35–e71

9. Sundt TM, Jneid H. Guideline update on indications for transcatheter aortic valve implantation based on the 2020 american college of cardiology/american heart association guidelines for management of valvular heart disease. JAMA cardiology. 2021;6:1088–1089

10. Lawton JS, Tamis-Holland JE, Bangalore S, Bates ER, Beckie TM, Bischoff JM, et al. 2021 acc/aha/scai guideline for coronary artery revascularization: A report of the american college of cardiology/american heart association joint committee on clinical practice guidelines. Circulation. 2022;145:e18–e114

11. Kristensen SD, Knuuti J, Saraste A, Anker S, Botker HE, Hert SD, et al. 2014 esc/esa guidelines on non-cardiac surgery: Cardiovascular assessment and management: The joint task force on non-cardiac surgery: Cardiovascular assessment and management of the european society of cardiology (esc) and the european society of anaesthesiology (esa). European heart journal. 2014;35:2383–2431

12. Gerhard-Herman MD, Gornik HL, Barrett C, Barshes NR, Corriere MA, Drachman DE, et al. 2016 aha/acc guideline on the management of patients with lower extremity peripheral artery disease: Executive summary: A report of the american college of cardiology/american heart association task force on clinical practice guidelines. Circulation. 2017;135:e686–e725

13. Powers WJ, Rabinstein AA, Ackerson T, Adeoye OM, Bambakidis NC, Becker K, et al. Guidelines for the early management of patients with acute ischemic stroke: 2019 update to the 2018 guidelines for the early management of acute ischemic stroke: A guideline for healthcare professionals from the american heart association/american stroke association. Stroke. 2019;50:e344–e418

14. Jadhav AP, Desai SM, Jovin TG. Indications for mechanical thrombectomy for acute ischemic stroke: Current guidelines and beyond. Neurology. 2021;97:S126–s136

15. Cook AM, Morgan Jones G, Hawryluk GWJ, Mailloux P, McLaughlin D, Papangelou A, et al. Guidelines for the acute treatment of cerebral edema in neurocritical care patients. Neurocritical care. 2020;32:647–666

16. Schizodimos T, Soulountsi V, Iasonidou C, Kapravelos N. An overview of management of intracranial hypertension in the intensive care unit. Journal of anesthesia. 2020;34:741–757

17. Massari D, de Keijzer IN, Scheeren TWL. Cerebral monitoring in surgical icu patients. Current opinion in critical care. 2021;27:701–708

18. Rohaut B, Eliseyev A, Claassen J. Uncovering consciousness in unresponsive icu patients: Technical, medical and ethical considerations. Critical Care. 2019;23:78

19. Boles J-M, Bion J, Connors A, Herridge M, Marsh B, Melot C, et al. Weaning from mechanical ventilation. European Respiratory Journal.29:1033–1056

20. Edelson DP, Churpek MM, Carey KA, Lin Z, Huang C, Siner JM, et al. Early warning scores with and without artificial intelligence. JAMA network open. 2024;7:e2438986–e2438986

21. Shillan D, Sterne JAC, Champneys A, Gibbison B. Use of machine learning to analyse routinely collected intensive care unit data: A systematic review. Critical Care. 2019;23:284

22. Samuels MA. The brain-heart connection. Circulation. 2007;116:77–84

23. Ahn S-H, Lee J-S, Yun M-S, Han J-H, Kim S-Y, Kim Y-H, et al. Explanatory power and prognostic implications of factors associated with troponin elevation in acute ischemic stroke. J Stroke. 2023;25:141–150

24. Ahn SH, Kim YH, Shin CH, Lee JS, Kim BJ, Kim YJ, et al. Cardiac vulnerability to cerebrogenic stress as a possible cause of troponin elevation in stroke. J Am Heart Assoc. 2016;5:e004135

25. Pinsky MR, Bedoya A, Bihorac A, Celi L, Churpek M, Economou-Zavlanos NJ, et al. Use of artificial intelligence in critical care: Opportunities and obstacles. Critical Care. 2024;28:113

26. Teasdale G, Maas A, Lecky F, Manley G, Stocchetti N, Murray G. The glasgow coma scale at 40 years: Standing the test of time. The Lancet. Neurology. 2014;13:844–854

27. Zietemann V, Georgakis MK, Dondaine T, Müller C, Mendyk AM, Kopczak A, et al. Early moca predicts long-term cognitive and functional outcome and mortality after stroke. Neurology. 2018;91:e1838–e1850

28. Hochreiter S, Schmidhuber J. Long short-term memory. Neural computation. 1997;9:1735–1780

29. Srivastava N, Hinton G, Krizhevsky A, Sutskever I, Salakhutdinov R. Dropout: A simple way to prevent neural networks from overfitting. J. Mach. Learn. Res. 2014;15:1929–1958

30. Kwon JM, Lee Y, Lee Y, Lee S, Park J. An algorithm based on deep learning for predicting in-hospital cardiac arrest. Journal of the American Heart Association. 2018;7

31. Subbe CP, Kruger M, Rutherford P, Gemmel L. Validation of a modified early warning score in medical admissions. QJM : monthly journal of the Association of Physicians. 2001;94:521–526

32. Lee CK, Samad M, Hofer I, Cannesson M, Baldi P. Development and validation of an interpretable neural network for prediction of postoperative in-hospital mortality. NPJ digital medicine. 2021;4:8

33. Xue B, Li D, Lu C, King CR, Wildes T, Avidan MS, et al. Use of machine learning to develop and evaluate models using preoperative and intraoperative data to identify risks of postoperative complications. JAMA network open. 2021;4:e212240

34. Zhang X, Fei N, Zhang X, Wang Q, Fang Z. Machine learning prediction models for postoperative stroke in elderly patients: Analyses of the mimic database. Frontiers in aging neuroscience. 2022;14:897611

35. Cho KJ, Kim JS, Lee DH, Lee SM, Song MJ, Lim SY, et al. Prospective, multicenter validation of the deep learning-based cardiac arrest risk management system for predicting in-hospital cardiac arrest or unplanned intensive care unit transfer in patients admitted to general wards. Critical care (London, England). 2023;27:346

36. Lundberg SM, Lee S-I. A unified approach to interpreting model predictions. Advances in neural information processing systems. 2017;30

37. Abgrall G, Holder AL, Chelly Dagdia Z, Zeitouni K, Monnet X. Should ai models be explainable to clinicians? Critical Care. 2024;28:301

38. Albers GW, Marks MP, Kemp S, Christensen S, Tsai JP, Ortega-Gutierrez S, et al. Thrombectomy for stroke at 6 to 16 hours with selection by perfusion imaging. N Engl J Med. 2018;378:708–718

39. Nogueira RG, Jadhav AP, Haussen DC, Bonafe A, Budzik RF, Bhuva P, et al. Thrombectomy 6 to 24 hours after stroke with a mismatch between deficit and infarct. N Engl J Med. 2018;378:11–21

40. Perkins GD, Callaway CW, Haywood K, Neumar RW, Lilja G, Rowland MJ, et al. Brain injury after cardiac arrest. *Lancet (London*, England). 2021;398:1269–1278

41. Esteva A, Robicquet A, Ramsundar B, Kuleshov V, DePristo M, Chou K, et al. A guide to deep learning in healthcare. Nature medicine. 2019;25:24–29

42. Sawyer RG, Leon CA. Common complications in the surgical intensive care unit. Critical care medicine. 2010;38:S483–493

43. Druedahl LC, Price WN, II, Minssen T, Sarpatwari A. Use of artificial intelligence in drug development. JAMA network open. 2024;7:e2414139–e2414139

